# Geospatial Modeling Study Assessing Population Level Accessibility to Medical College Hospitals in India

**DOI:** 10.1101/2024.08.11.24311839

**Authors:** Harsh Thakkar, Chaitanya Reddy, Varun Raj Passi, Aamir Miyajiwala, Sahil Kale, Ankit Raj, Siddhesh Zadey

## Abstract

**Background:** While studies have investigated the availability of Medical College Hospitals (MCHs) in India, data on geographic accessibility are limited. Our study examines current geographic accessibility to these MCHs across 36 states and union territories and 735 districts.

**Methods:** We validated MCH data from the National Health Profile 2022, using travel-time rasters from the Malaria Atlas Project (2019) and population data from WorldPop (2020). We examined the density of MCHs per million population and the median travel time to the nearest MCH. We assessed the Access Population Coverage (APC), defined as the proportion of the population within 30, 60, 90, and 120 minutes by motorized transport, and within 30 and 60 minutes by walking, to the nearest MCH.

**Results:** In 2022, India had an average density of 0.47 MCHs per million. The median travel time to the nearest MCH was 67.94 minutes by motorized transport and 589.82 minutes by walking. 71.76% of the population could access the nearest MCH within 60 minutes by motorized transport. The APC was 64.20% within 60 minutes by motorized transport in rural areas vs. 92.34% in urban areas. The APC within 60 minutes by motorized transport for public MCHs was 63.62%, while that for private was 45.95%.

**Conclusions:** Median travel time and APC help assess geographic accessibility. Our study found wide disparities in MCH access across Indian states and rural vs. urban areas. These analyses can guide the optimal placement of new MCHs.

## 1. Background

Medical college hospitals (MCH) play an essential role in healthcare systems worldwide. They provide hands-on clinical training to medical students, act as centers of research setting standards of care, (1) and offer specialized clinical care to patients. In lower-middle-income countries (LMICs) MCHs often act as the last points of referral, providing the highest level of specialized care available. (2)

India was estimated to have 648 MCHs in 2022. (3) MCHs are essential in providing subsidized or free tertiary care services, as a large proportion of them (estimated to be 53.39% in 2022) are publicly owned. Privately owned MCHs often provide subsidized care in exchange for government concessions. They play a crucial role in emergencies like trauma, maternal and newborn care, and non-communicable diseases, as public tertiary centers have limited capacity. MCHs also improve local primary health care in their surrounding region by providing outreach services. (4) Therefore, MCHs act as essential stakeholders in the healthcare system of their geographical area. Provinces with higher MCH density in India perform better on major health indicators, such as the maternal mortality ratio (MMR), infant mortality rate (IMR), and the percentage of institutional deliveries. (5)

Given the importance of MCHs in India’s public health system, it is important to study their growth and distribution in the country. There has been a rapid growth in the total number of MCHs in India in the last few decades. An important feature of this growth is the increasing involvement of the private sector in medical education. (4) However, the distribution and growth of MCHs are not even. MCHs are heavily clustered in provincial capitals and major cities that already have established and well-functioning MCHs. Out of 207 newly established MCHs between 2009 and 2019, 139 (67.15%) were located within 50km of an old MCH. This unequal distribution of MCHs is starker for private MCHs, with 60.6% of all private MCHs in the country located in a handful of provinces in the southern part of India, i.e., Maharashtra, Andhra Pradesh, Telangana, Karnataka, Kerala, Tamil Nadu, and Puducherry. (5)

Availability refers to the extent to which resources are available to meet patients’ needs. The number of health facilities per defined population size is referred to as the health facility density and is a measure of the availability of care. Geographic accessibility refers to the proximity to care, with an element of time, and delineates how easily patients can reach the provider’s location. (6) While studies have mapped the distribution and density of MCHs in India, (4,5) geographic accessibility in terms of time and the population covered remains to be studied. Timely access to tertiary healthcare services is necessary for optimal outcomes, particularly in time-sensitive emergencies such as stroke, (7) acute coronary syndromes, (8) surgical, and maternal and child health emergencies. Poor access to specialized care also increases costs due to travel and longer lost wages. (9–11) Consequently, geographic accessibility to MCHs constitutes an important component of moral capital in the country. (12)

We evaluated geographic accessibility to MCHs using density per million people, travel time to the nearest MCH, and access population coverage (APC). For travel time and APC, two modes of transport were considered: motorized and walking. These modes correspond to the two travel-time friction-surface rasters available through the Malaria Atlas Project (MAP),(13) which is one of the most widely validated and frequently used global datasets for geospatial accessibility modelling. Motorized and walking surfaces capture two extremes of mobility in the Indian context: (i) the dominant mode with optimal speeds for medical travel (motorized), and (ii) a lowest mobility scenario (walking) to reflect populations without transport access, especially in rural and remote regions. These rasters incorporate terrain, slope, land cover, and road networks, which makes them suitable for nationwide modelling. Estimates were provided for 735 districts across 36 Indian states and union territories. We also analyzed disparities between rural and urban areas and public versus private hospitals.

## 2. Methods

### 2.1 Data Sources

To comprehensively map MCH densities and their travel times, the study required the collection, extraction, and collation of data from the National Health Profile (NHP) Report 2022 published by the Central Bureau of Health Intelligence. (3) The report lists active MCHs, including addresses, ownership, beds, and training intake. Government-owned MCHs and those jointly managed by the government and non-profits were classified as public; all others were classified as private. Population data came from the 2020 WorldPop, providing high-resolution (1 km²) UN-adjusted counts for India. (14) Motorized and walking friction rasters (1 km²) were sourced from the 2019 Malaria Atlas Project (MAP). MAP, an international scientific initiative, focuses on mapping global malaria responses. (13) Friction rasters are informational maps that show how much it costs to go through them, usually expressed in time units. (15) These friction rasters account for several physical and environmental variables that may impede or prolong travel. The administrative borders of India were drawn from the publicly available shapefile. (4,16) A global raster with ordinal catchment area (CA) categories based on population densities and proximity to high-density urban areas provided standardized rural-urban regions. (17) We classified CA categories >7 as rural.

### 2.2 Outcomes

We examined three primary outcomes: MCH density, measured as the number of MCHs per million people to assess geographic distribution; travel time to the nearest MCH, reporting median and interquartile range (IQR) values; and Access Population Coverage (APC), defined as the percentage of the population with timely access to the nearest MCH. Timely access thresholds were set at 30, 60, 90, and 120 minutes for motorized transport and 30 and 60 minutes for walking, combining population and timeliness aspects to evaluate accessibility. (18–21)

### 2.3 Primary Analysis

Before geocoding, MCH addresses were cleaned for machine readability by standardizing formats and correcting errors. Geocoding used the Google Maps API to select the best match for each address. Coordinates were checked for duplicates and implausible locations outside India.

Population data were obtained from WorldPop 2020 and rural-urban CA rasters. The global CA raster (1 km² resolution) was clipped to India’s boundary and split into rural (CA >7) and urban (CA ≤7) layers. WorldPop 2020 data were overlaid on these layers and resampled to align. Rural populations were weighted by 1, urban by ‘NA’, and the totals were summed. National, state, and district populations were aggregated using administrative boundaries.

Travel times were calculated using the Dijkstra algorithm on MAP friction surfaces, estimating the shortest time from each 1 km² pixel to the nearest MCH. Walking and motorized transport rasters provided travel times in minutes. Median and IQR values were computed at various levels. APCs used thresholds of 120 minutes (motorized) and 30 minutes (walking) in the primary analysis. Accessibility rasters were dichotomized, matched to population rasters, and used to calculate timely access proportions.

**Figure 1** describes the analysis pipeline, using rural motorized APC estimation as an example. We have previously discussed the details in our research papers and used them to investigate access to surgical facilities in India. (20–25)

**Figure 1:**
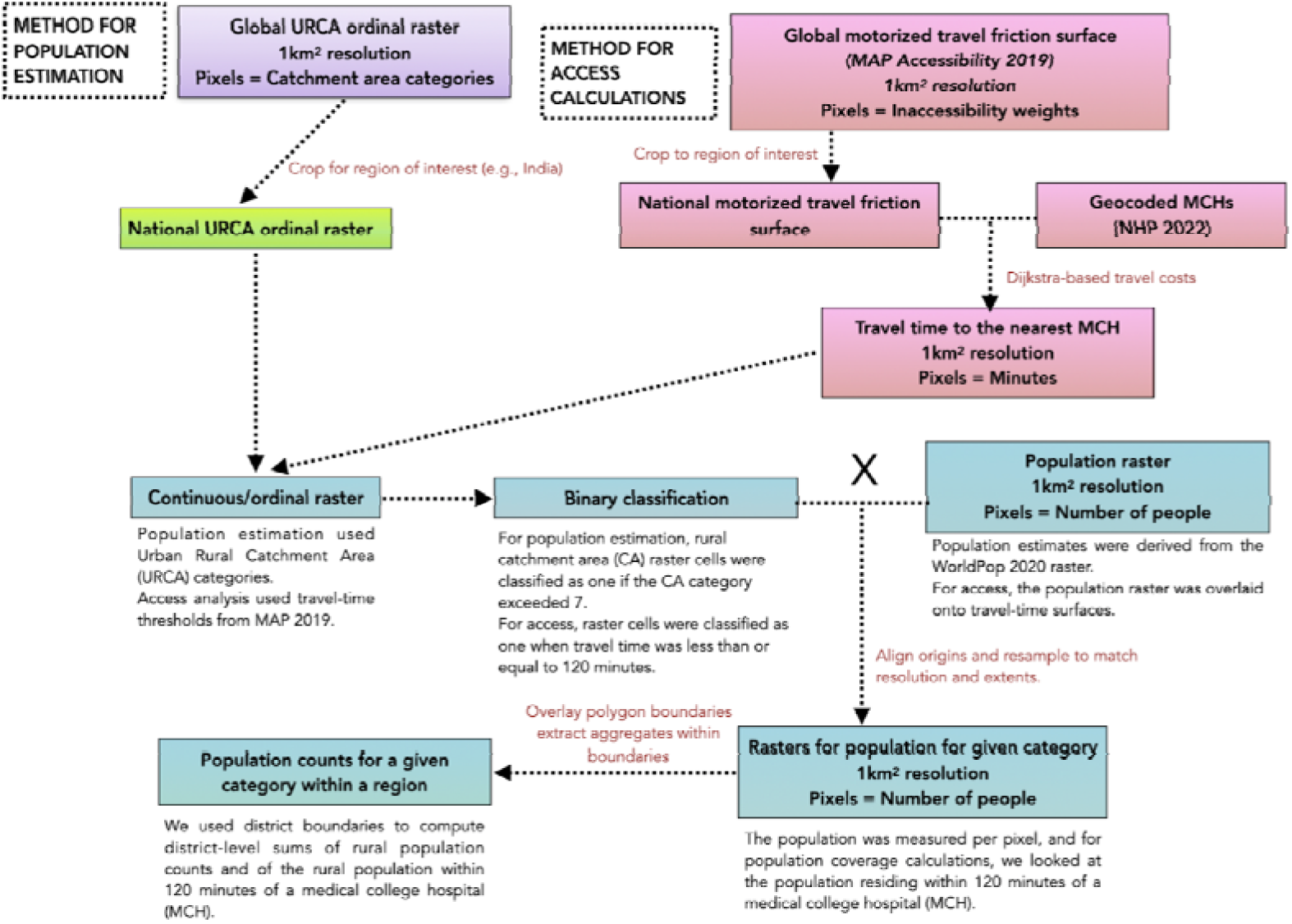
Raster-based estimation pipeline used for creating criteria-specific regional population counts. Here, we focus on rural motorized APC as an example.

### 2.4 Sensitivity Analyses

To assess the robustness of our primary estimates, we conducted a sensitivity analysis using multiple APC scenarios. First, APC was calculated assuming motorized transportation for time thresholds of 30, 60, and 90 minutes. Recognizing that a substantial share of the population may lack access to motorized transport, we included APC when walking was the mode of travel, applying 60 minute thresholds. This approach allowed us to examine how assumptions about travel mode and time thresholds influence APC estimates and to evaluate the stability of our findings across alternative accessibility scenarios.

### 2.5 Correlation Analysis with District-Level Universal Health Coverage (UHC) Index

We conducted a district-level correlation analysis to examine the relationship between geographic accessibility to MCHs, measured by APC values, and the district-level Universal Health Coverage (UHC) index (24), which served as the reference indicator. We chose the UHC index since it is the relevant indicator of overall health system performance. To ensure comparability, districts for which UHC index values were not reported were excluded, resulting in a final sample (n) of 687 districts. This analysis was undertaken to assess whether variations in geographic access to MCH are aligned with broader dimensions of health service coverage and system strength captured by the UHC index. APC estimates were matched to corresponding district-level UHC index values using name-based district identifiers. This ecological exploratory association analysis used the nonparametric Spearman rank correlation coefficient (denoted by ‘R’), given the nonnormal data distribution. A p-value of <0.05 was considered statistically significant.

## 3. Results

### 3.1 MCH Density in India

According to NHP 2022, India had 648 MCHs. Tamil Nadu had the highest number of MCHs (71), followed by Karnataka (67) and Uttar Pradesh (67). Nagaland, Ladakh and Lakshadweep had no MCHs. The national density was 0.47 MCHs per million people **(eFigure 1A)**.

Twenty-three states/union territories exceeded the national MCH density average. Puducherry had the highest density (6.47), while Bihar had the lowest (0.16). Among 735 districts, 359 lacked MCHs. Puducherry also led in district-level density (7.35 per million). Only 104 districts (14.14%) had a density above one, while 264 (35.91%) surpassed the national average. **(eFigure 1B)**.

In rural areas, 210 (32.41%) MCHs were located. Seven states (19.44%) had no rural MCHs per million people, while eight (22.22%) surpassed the national average. Puducherry had the highest rural MCH density at 9.91. In urban areas, three states (8.33%) had no MCHs per million people. However, 29 states (80.55%) had densities above the national average. Sikkim had the highest urban MCH density at 20.71.

Of the 648 MCHs, 346 (53.40%) were public, and 302 (46.60%) were private. Sikkim had no public MCHs, but 11 states (30.55%) had public MCH densities above the national average. The Andaman and Nicobar Islands had the highest public MCH density at 2.76.

Seven states (19.44%) had no private MCHs, while five states (13.88%) exceeded the national density. Puducherry had the highest private MCH density at 5.03.

### 3.2 Travel time to the nearest MCH

#### 3.2.1 Motorized Transport

The national median (Interquartile Range or IQR) travel time to the nearest MCH by motorized transport was 67.94 (41.32, 110.42) minutes **(eFigure 2A)**. Seventeen states/union territories reported shorter travel times than the national value. Access was poor in Ladakh, where the median travel time exceeded 591.74 minutes. Five states had median travel times under 30 minutes, and 17 under 60 minutes. At the district level, 60.81% districts had median travel times below the national median value. Mumbai in Maharashtra reported the shortest median travel time of 1.89 (1.16, 2.67) minutes, whereas Anjaw in Arunachal Pradesh had the longest travel time of 1576.80 (1136.23, 2054.06) minutes **(eFigure 2B)**. Rural areas had a median travel time of 69.04 (42.54, 111.38) minutes compared with 25.92 (8.79, 49.47) minutes in urban areas. Seventeen rural and 33 urban states/union territories reported median travel times shorter than the national median value **(eFigure 2C)**.

**Figure 2:**
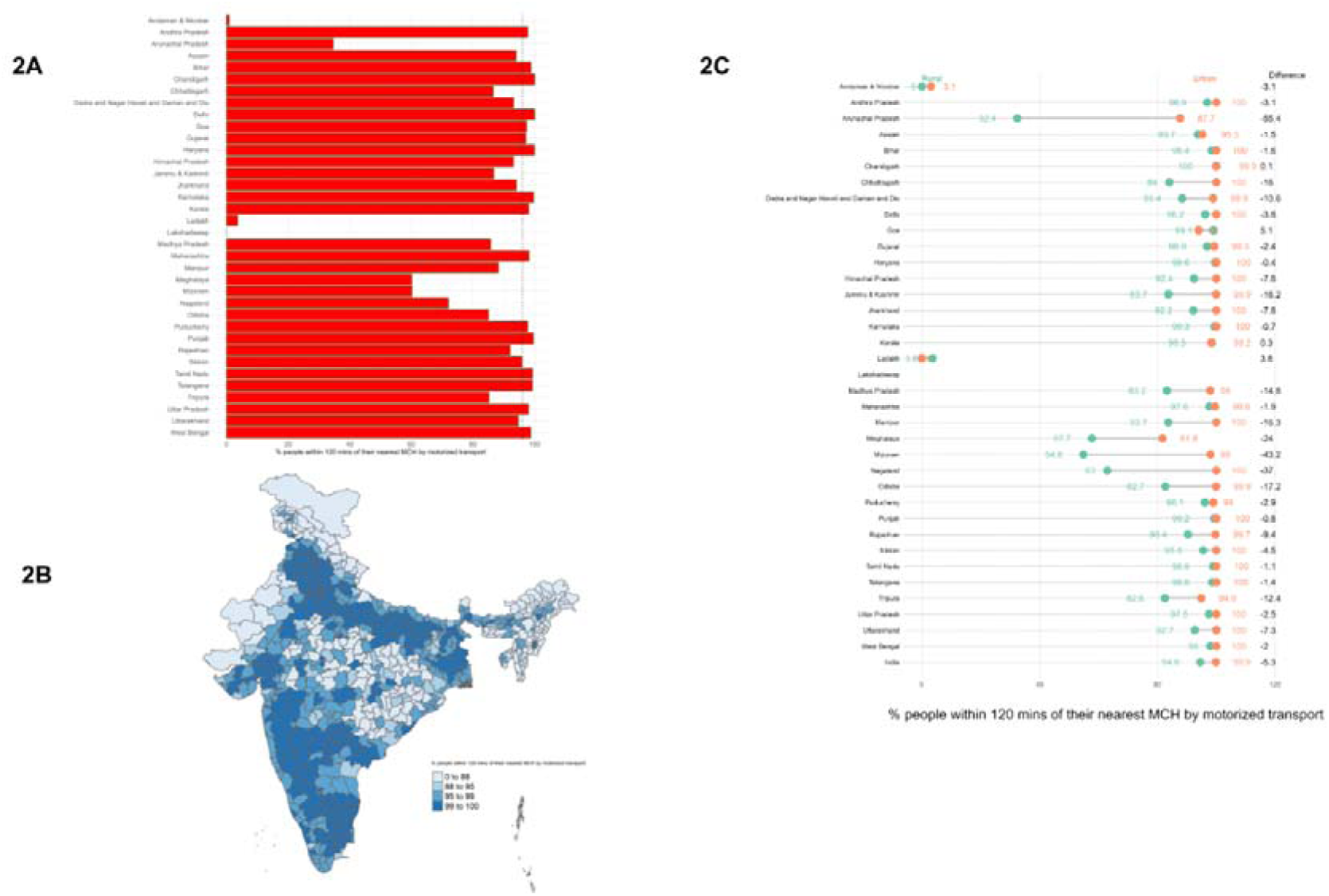
APC values to access the nearest MCH within 120 minutes by motorized transport for A. States and union territories with the national value of 95.94% (black dotted line). B. Districts and C. Rural and urban areas. The difference is shown as the rural APC minus the urban APC, in percentage points.

The median travel time to public MCHs was 77.46 (47.86, 120.38) minutes **(eFigure 3A)**, compared with 106.69 (61.67, 181.94) minutes for private MCHs **(eFigure 3B)**. Chandigarh reported the shortest median travel time to public MCH at 6.33 minutes (4.16, 7.93), compared with Ladakh, which reported the longest median travel time at 591.74 minutes. For private MCHs, travel time was shortest in Puducherry at 8.25 (4.70, 12.23) minutes and longest in Arunachal Pradesh at 856.29 minutes. At the district level, median travel times for public MCHs ranged from 1.89 minutes in Mumbai to 1576.80 minutes in Anjaw **(eFigure 3C)**. Similarly, for private MCHs, median travel times ranged from 5.69 minutes in Hyderabad to 1895.79 minutes in Anjaw, highlighting severe disparities **(eFigure 3D)**.

**Figure 3:**
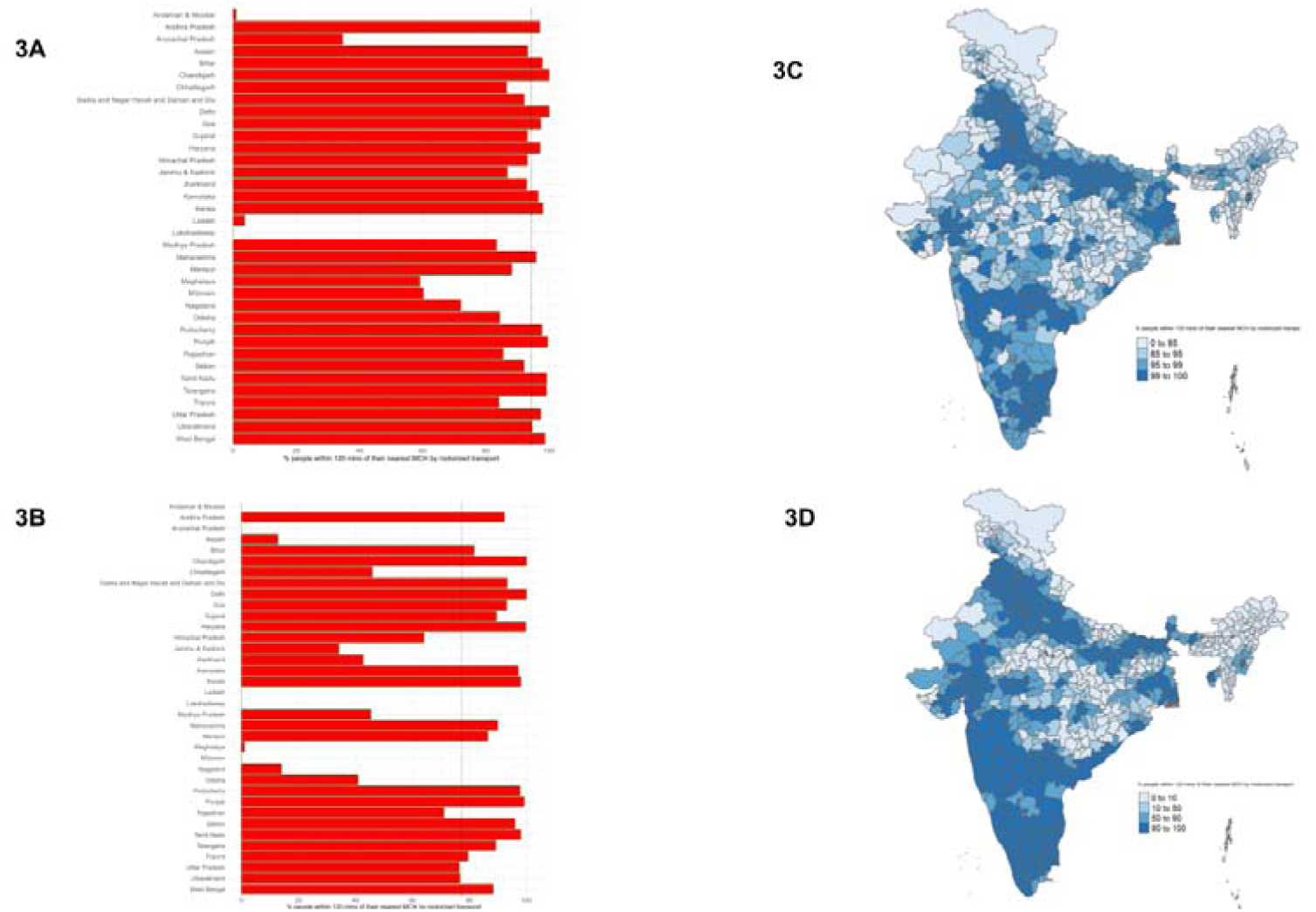
**APC values to access the nearest MCH within 120 minutes by motorized transport for A. States and union territories with public MCHs. B. States and union territories with private MCHs. C. Districts with public MCHs and D. Districts with private MCHs.**

#### 3.2.2 Walking

The national median travel time (IQR) to the nearest MCH by walking was 589.82 (359.49, 913.74) minutes. Eighteen states/union territories had shorter travel times, and 17 had longer travel times than the national median value. Lakshadweep had no median travel time value because there were no MCHs. Chandigarh reported the shortest travel time at 58.60 (39.14, 75.13) minutes, whereas Ladakh reported the longest at 4120.86 (3353.96, 4910.96) minutes **(eFigure 4A)**. At the district level, 60.95% reported a median travel time below the national median value. Mumbai had the shortest travel time of 22.13 (11.06, 30.55) minutes, while Leh recorded the longest at 4466.33 (3786.09, 5176.35) minutes **(eFigure 4B)**. The rural population experienced substantially longer travel time by walking with a median travel time of 597.18 (368.41, 919.30) minutes, as compared to the urban value of 269.57 (93.13, 509.30) minutes. Among rural areas, the population residing in Chandigarh had the shortest median travel time of 59.65 (48.84, 92.89) minutes, while among urban areas, the population residing in Sikkim had the shortest median travel time of 48.34 (25.78, 215.33) minutes **(eFigure 4C)**.

**Figure 4:**
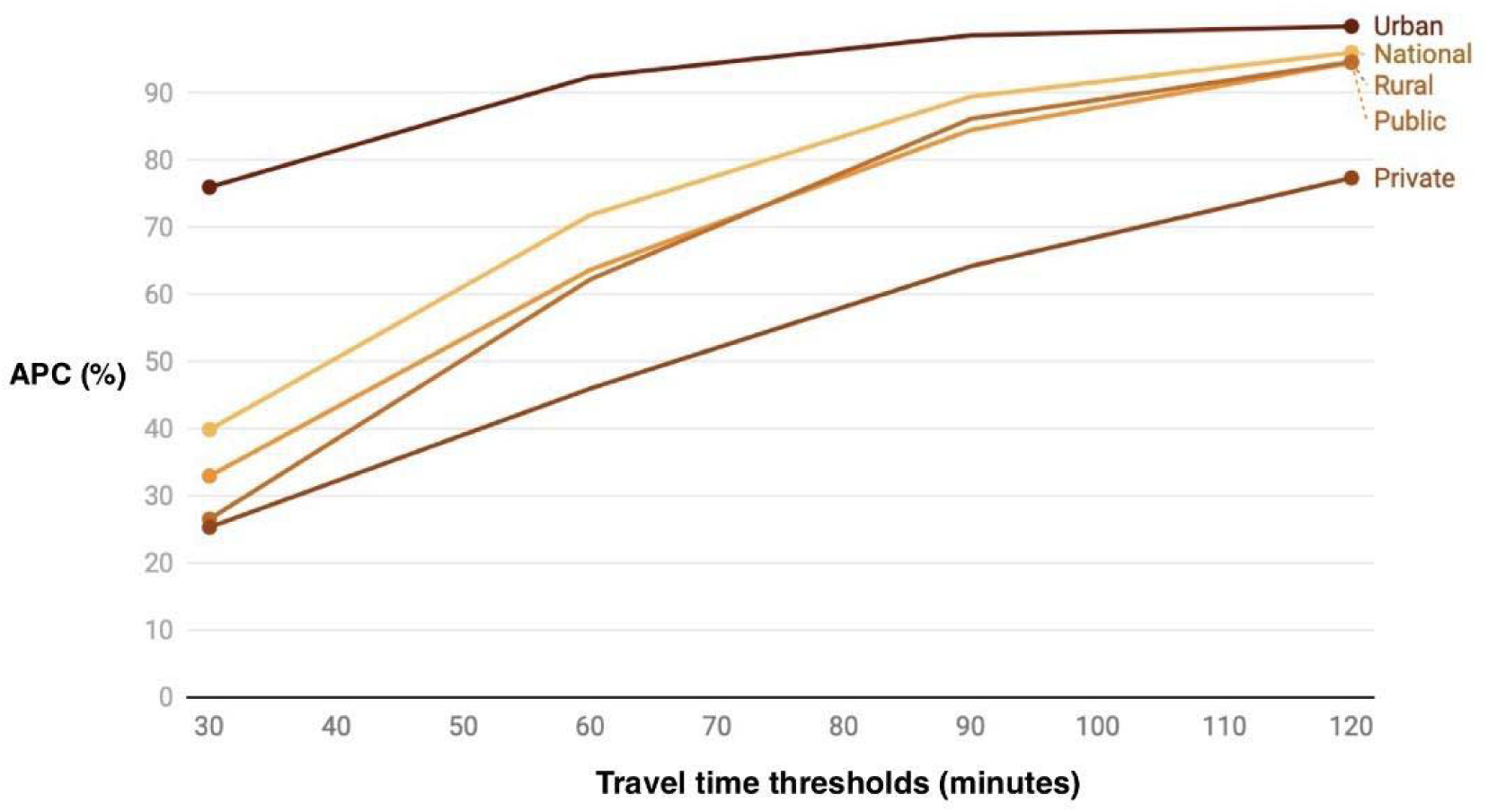
Sensitivity analyses for motorized transport APC values at different time thresholds (30, 60, 90 and 120 minutes) across various areas (rural and urban) and sectors (public and private).

For public MCHs, the national median travel time, by walking, was 687.91 (426.49, 1044.83) minutes **(eFigure 5A)**, increasing to 992.37 (567.88, 1672.34) minutes for private MCHs **(eFigure 5B)**. Chandigarh reported the shortest public MCH time at 58.60 (39.14, 75.13) minutes, while Ladakh had the longest at 4120.86 (3353.96, 4910.96) minutes. For private MCHs, Puducherry had the shortest median travel time at 90.82 (53.40, 132.16) minutes, and Arunachal Pradesh had the longest at 5853.52 (4964.69, 6905.61) minutes. District-level median travel time ranged from 22.13 (15.27, 30.55) minutes in Mumbai for public MCHs **(eFigure 5C)** to 7322.25 (6815.12, 7973.86) minutes in Upper Dibang Valley for private MCHs **(eFigure 5D)**.

**Figure 5:**
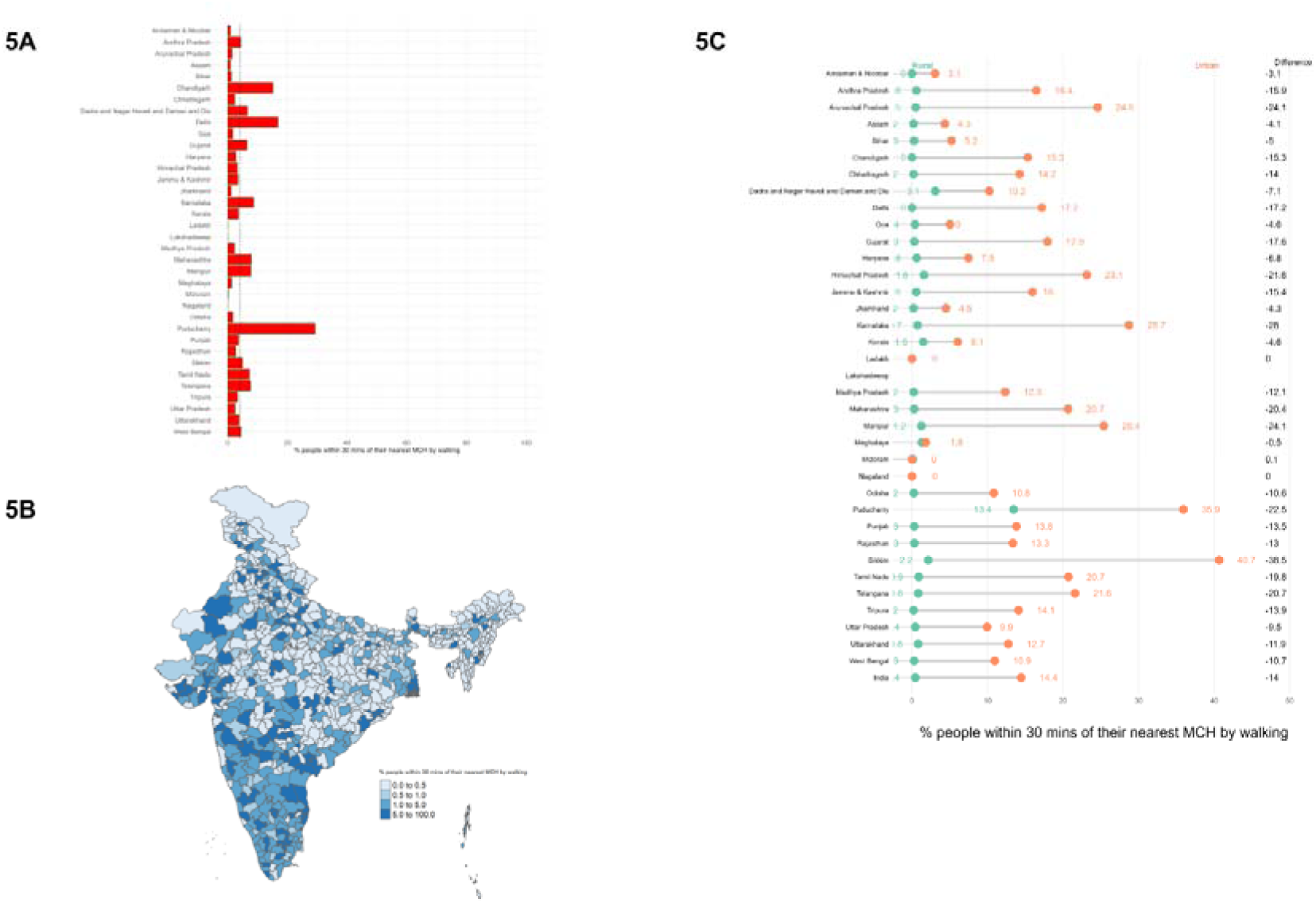
APC values to access the nearest MCH within 30 minutes by walking for A. States and union territories with the national value of 4.22% (black dotted line). B. Districts and C. Rural and urban areas. The difference is shown as the rural APC minus the urban APC, in percentage points.

### 3.3 Access population coverage (APC)

#### 3.3.1 Motorized transport

By motorized transport, timely access to an MCH was defined as reaching a hospital within 120 minutes. The national APC was 95.94%. Lakshadweep had no MCH access within 120 minutes. In the Andaman and Nicobar Islands, under 1% of the population could reach an MCH within 120 minutes. In Chandigarh and Delhi, 100% of the population was within 120 minutes of an MCH **(Figure 2A)**. Fourteen (1.90%) districts had no access to an MCH. Less than 1% of the population in three districts and 100% of the population in 112 (15.23%) districts had timely access. A total of 307 (41.76%) districts had APCs above 99% **(Figure 2B)**.

Nationally, 94.55% of the rural population and 99.86% of the urban population had access to an MCH (difference is -5.31 percentage points). Rural areas in Lakshadweep, Andaman and Nicobar Islands, and urban areas in Ladakh had zero APCs. Chandigarh had the highest rural APC (100%), while Ladakh had the lowest (3.60%). Urban areas in 17 states/union territories had 100% APC, while Andaman and Nicobar Islands had the lowest (3.05%). Arunachal Pradesh had the largest rural-urban APC gap (-55.4%), and Chandigarh had the smallest (0.1%) **(Figure 2C)**.

Nationwide, 94.46% of the population had access to public MCHs **(Figure 3A)**, while 77.29% could reach private MCHs within 120 minutes **(Figure 3B)**. Lakshadweep had no public MCH access and four states/union territories had no private MCH access. Chandigarh and Delhi had 100% APC for both types of MCHs. At the district level, 15 (2.04%) districts lacked public MCH access, and 134 (18.23%) districts had no private MCHs. Ninety-seven (13.19%) districts with public MCHs **(Figure 3C)** and 81 (11.02%) districts with private MCHs achieved 100% APC **(Figure 3D)**. Compared to the national APC, 455 (61.90%) districts had higher public MCH access, and 412 (56.05%) districts had higher private MCH access.

##### 3.3.1.1 Sensitivity analyses

Sensitivity analyses for 30, 60, and 90 minute motorized thresholds showed persistent disparities **(Figure 4)**. Within 30 minutes, national APC was 39.85%, with urban (75.92%) far exceeding rural (26.54%), a 49.38-point gap. Eleven states/union territories exceeded 50% APC, while Ladakh and Lakshadweep had none. Public and private MCH had an APC of 32.95% and 25.32%, respectively. At 60 minutes, APC rose to 71.76%, with urban (92.34%) exceeding rural (62.20%) by 30.14 points. Sixteen states/union territories surpassed 75% APC, though Ladakh and Andaman and Nicobar Islands lagged. By 90 minutes, APC reached 89.40%, with urban at 98.51% and rural at 86.10%. Despite improved access, rural and remote areas still faced major challenges.

##### 3.3.1.2 Association with UHC Index

In the correlation analysis with UHC index, the association was strongest for access within 30 minutes with a Spearman Rank correlation (R) of 0.308 ([95% CI: 0.239, 0.374], n=687, p <0.001) **(eFigure 6A).** It remained positive at 60 minutes (R = 0.275 [95% CI: 0.205, 0.343], n=687, p <0.001) **(eFigure 6B)** 90 minutes (R = 0.218 [95% CI: 0.146, 0.288], n=687, p <0.001) **(eFigure 6C)** and 120 minutes (R = 0.125 [95% CI: 0.050, 0.198], n=687, p=0.001) (eFigure 6D).

**Figure 6:**
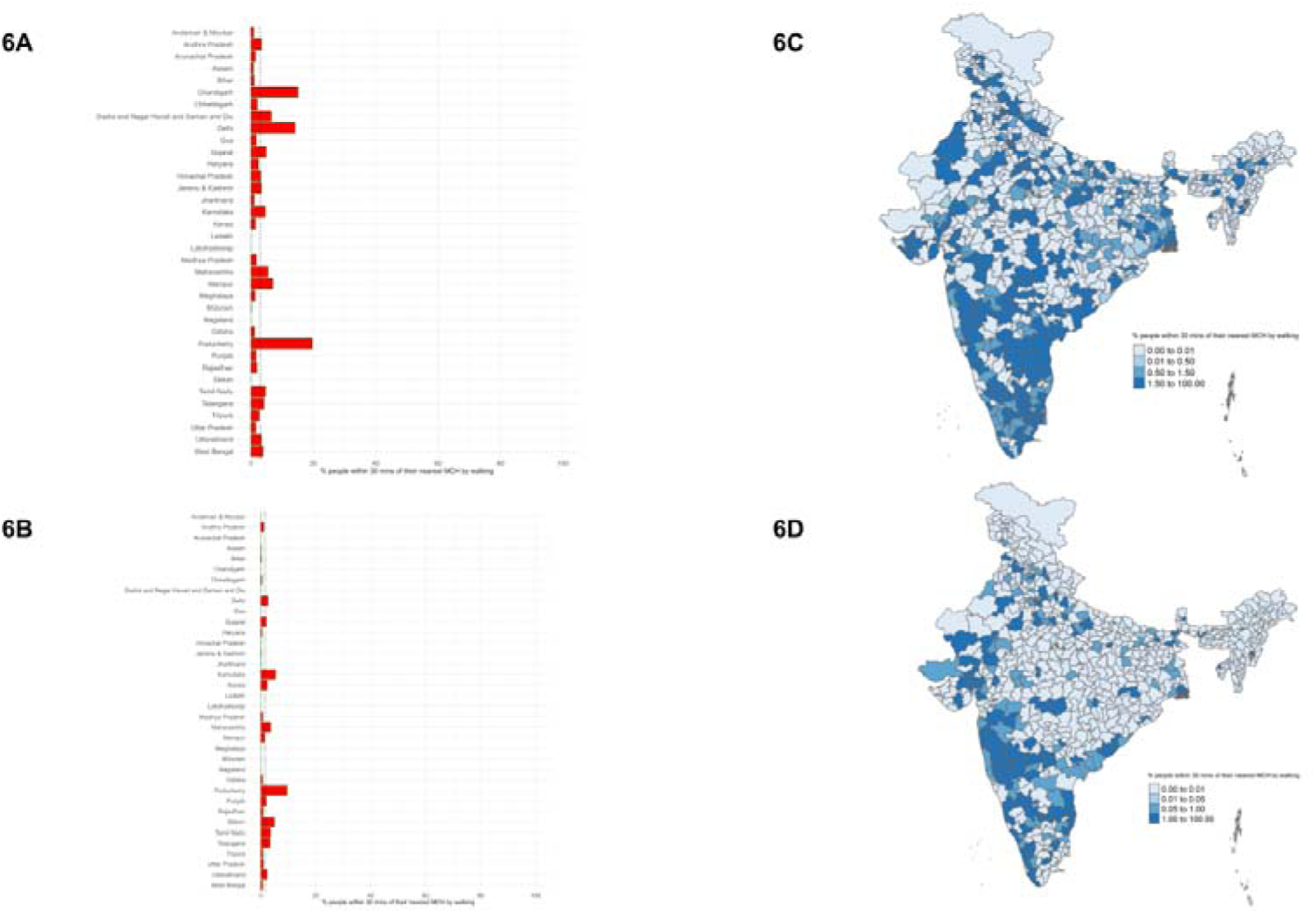
APC values to access the nearest MCH within 30 minutes by walking for A. States and union territories with public MCHs. B. States and union territories with private MCHs. C. Districts with public MCHs and D. Districts with private MCHs.

#### 3.3.2 Walking

By walking, timely access to an MCH meant reaching it within 30 minutes. Nationally, only 4.22% of the population had timely access. Lakshadweep, Ladakh and Nagaland had no access, while Puducherry (29.24%) and Delhi (16.80%) had the highest APC values. Only three states/union territories exceeded 10% APC **(Figure 5A)**. At the district level, 350 (47.61%) districts had zero APC. Mumbai had the highest APC (71.86%), while Kullu had an APC below 1% **(Figure 5B)**. Nationally, rural access (less than 1%) lagged far behind urban access (14.44%). Puducherry led in rural APC (13.43%), and Sikkim led in urban APC (40.69%). Sikkim also had the largest rural-urban gap (-38.5%), while Mizoram had the smallest (0.1%) **(Figure 5C)**.

For public MCHs, 2.96% of the population had timely access **(Figure 6A)**, compared to 1.53% for private MCHs **(Figure 6B)**. Four states/union territories had no access to public MCHs, whereas ten had no access to private MCHs. Puducherry led in both public (19.68%) and private (9.56%) APC. At the district level, 425 (57.82%) districts had no access to public MCHs, and 550 (74.82%) districts had no access to private MCHs. Mumbai had the highest public APC (71.83%) **(Figure 6C)**, and Chengalpattu led in private APC (20.96%) **(Figure 6D)**. Compared to the national APC, 139 (18.91%) districts had higher APC for public MCHs, and 93 (12.65%) districts had higher APC for private MCHs.

##### 3.3.2.1 Sensitivity Analyses

Sensitivity analysis for the 60-minute walking threshold **(Figure 7)** showed 10.96% of the population had timely access. Chandigarh (68.46%) and Puducherry (65.75%) had the highest APC, while rural access (1.98%) lagged far behind urban access (35.27%). Only 1.90% of districts exceeded 50% APC, with Shahdara in Delhi leading at 99.80%. Access to public (7.95%) and private (4.86%) MCHs was limited, with 56.19% of districts lacking public MCHs and 73.19% lacking private MCHs. Shahdara in Delhi (99.80%) and Bangalore in Karnataka (51.81%) led in public and private access, respectively, with urban areas consistently outperforming rural areas.

**Figure 7:**
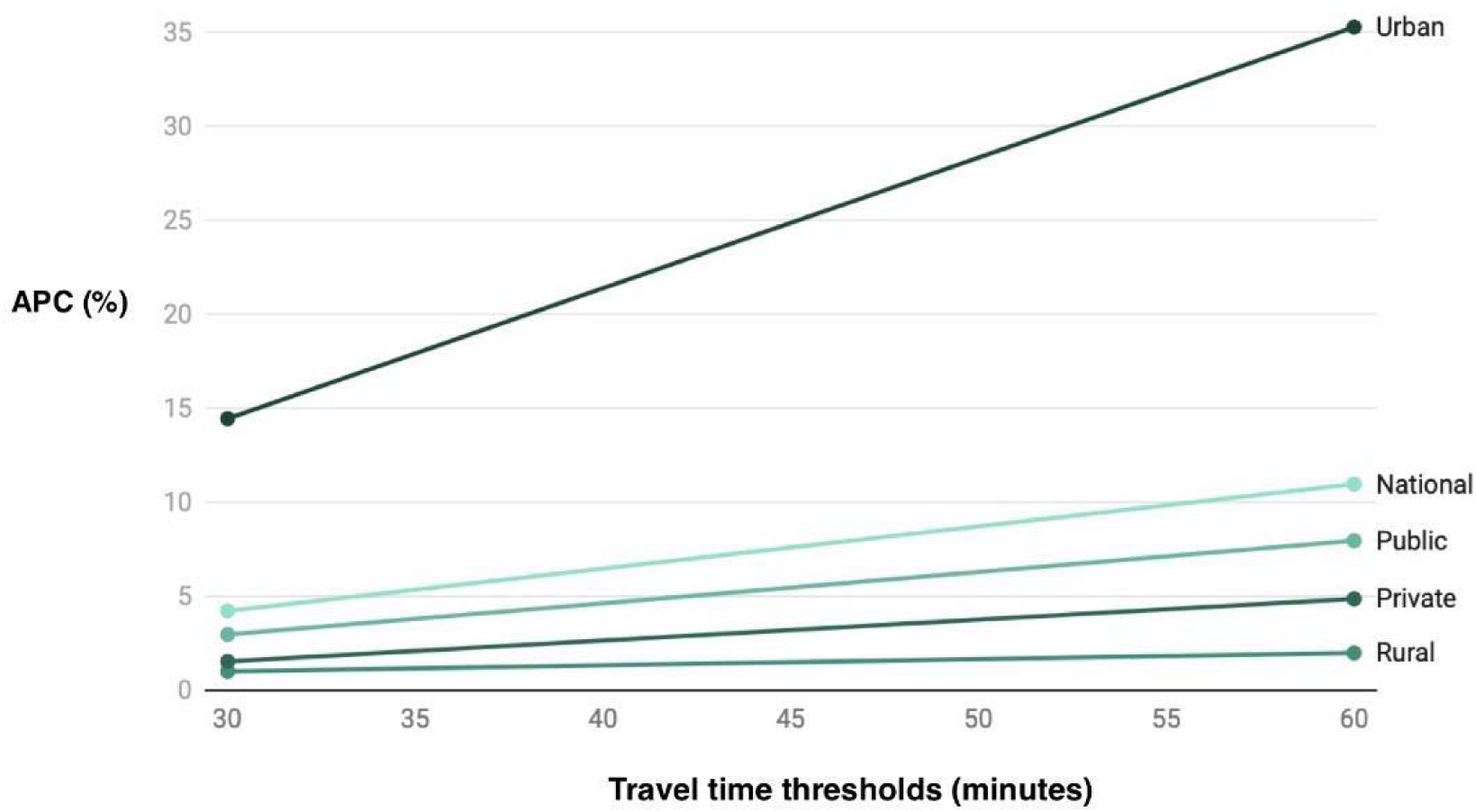
Sensitivity analyses for walking APC values at different time thresholds (30 and 60 minutes) across various areas (rural and urban) and sectors (public and private).

##### 3.3.2.2 Association with UHC Index

From correlational analysis, districts where a higher proportion of the population could reach MCHs by walking within shorter time limits tended to have better UHC index scores, with an R = 0.297, 95% CI: 0.228, 0.363, n=687, p <0.001 for 30 minutes **(eFigure 7A)** and R = 0.316, 95% CI: 0.248, 0.381, n=687, p <0.001 for 60 minutes **(eFigure 7B)**.

## 4. Discussion

Using the NHP 2022 report, we assessed MCH densities per million, median travel times (motorized and walking) and APC up to the district level in India. Nationally, India had 0.47 MCHs per million. Median travel times were 67.94 minutes (motorized) and 589.82 minutes (walking). By motorized transport, APC was 39.85% (30 minutes), 71.76% (60 minutes), 89.40% (90 minutes), and 95.94% (120 minutes). By walking, APC was 4.22% (30 minutes) and 10.96% (60 minutes). We observed a consistent positive association with APC by motorized transport across all travel time thresholds.

The observed positive direction and statistical significance of the correlations provide preliminary evidence for the expected relationship between APC and the UHC index. Districts with higher APC values reported higher UHC index values. The association between APC and UHC index was comparable between motorized transport and walking. The strongest association was seen for the time threshold of 30 minutes by walking . This finding is important because walking access reflects local service availability and is less influenced by transport infrastructure. The analysis did not account for confounding factors that could influence both ease of access and health system performance. The strength of the association also differed across various districts within the same state.

MCH density rose from 0.41 to 0.47 per million since 2019. Puducherry had the highest density, while Nagaland and Ladakh had zero MCHs, unchanged from 2019. (5) Our study aligns with international findings. Bangladesh has 0.44 teaching hospitals per million, similar to India’s 0.47. District disparities are significant, with travel times to the nearest tertiary hospital ranging from 30 minutes to 276 minutes. (26,27) The significant disparities in availability and accessibility between rural and urban areas are also consistent. (27) For example, Dhaka, Bangladesh’s capital, hosts 16 (45.71%) of the country’s 35 tertiary health centers. Similarly, 92.5% of Sub-Saharan Africa’s population lives within two hours of a hospital offering essential surgical services, comparable to India’s 95.94% accessibility by motorized transport. (28) Therefore, the patterns and values observed are similar to those of South Asian countries and other LMICs.

Density shows facility distribution but not accessibility. Our study is the first to calculate median travel times and APC for MCHs in India. Travel time depends on roads, terrain, and traffic, not just distance. Geographic features such as rivers and mountains also affect movement. Recent global modelling work shows that terrain ruggedness, slope and elevation can substantially increase travel time even where distances are short. (29) Hydrological barriers such as rivers, wetlands, and coastal inlets also create major discontinuities in travel networks, often requiring long detours or slower modes of transport. (17) In low- and middle-income countries where road networks are heterogeneous, these geographic features produce large variations in travel time to tertiary facilities. (27,28) Therefore, accessibility is better captured through travel-time modelling than density measures, because friction surfaces integrate these real-world constraints. APC, which incorporates the population distribution and the time required to reach the nearest MCH, provides a more meaningful indicator of accessibility and should guide the strategic placement of new MCHs.

Several states/union territories need policy attention. Ladakh has the longest travel times. In Andaman and Nicobar, less than 1% could reach an MCH within 120 minutes. Northeastern states also have poor access, with APCs far below the 71.6% national average. Severe rural-urban disparities exist. Rural areas had a median travel time of 69.04 minutes, compared to 25.92 minutes in urban areas. Only 62.20% of rural residents could reach an MCH within 60 minutes, versus 92.34% in urban areas. Given that 68.8% of India’s population is rural, (30) this gap is alarming. Limited access to specialized care worsens outcomes, especially in emergencies, and increases the reliance on unqualified providers. (31)

Rural-urban access gaps may result from poorer rural road infrastructure. In 2018-19, urban road density was 5,296.3 per 1,000 km², compared to 1,458.1 in rural areas. Growth of Private MCHs may have also contributed to this gap, as private MCHs, reliant on student fees and patient revenue, are concentrated in wealthier districts. (32,4) The per-capita net value added was estimated at INR 40925 and 98435 in rural and urban areas, respectively, in 2011. (33) Urban populations’ higher paying capacity attracts private MCHs. Better urban infrastructure also draws more private investment. (4) Additionally, health professionals in India prefer urban areas for better amenities and career growth. This helps city-based MCHs meet NMC faculty requirements more easily. (5)

Public MCHs are more accessible (63.62%) than private MCHs (45.95%), partly because they have more facilities (346 vs. 302). Private MCHs tend to be in wealthier districts, where unregulated placement may not align with healthcare needs, further affecting accessibility. (4)

The strategic placement of new MCHs can address disparities in geographic accessibility to MCHs. The use of location-allocation models (LAMs) can help policymakers locate MCHs at optimal locations, thereby improving accessibility. (34) Plans such as the central government’s proposal to convert district hospitals into MCHs (34), particularly in underserved districts, can also reduce disparities. Improving roads in rural areas through schemes such as the Pradhan Mantri Gram Sadak Yojana (PMGSY) can also enhance accessibility. (35) It has also been noted that alumni of MCHs located in rural areas are more inclined towards rural practice. (5) Following the World Health Organization’s recommendation of establishing MCHs away from major cities can increase the retention of doctors in rural areas, improving accessibility. (35)

### Limitations and Strengths

The study has limitations but key strengths. It relied on assumptions from source data and excluded vehicle ownership, facility quality, and non-land transport. Demographic-specific access was not analyzed. We could not analyse for specific transport modes such as private motorized transport, intermediate public transport or two-wheelers because validated national raster data were not available. As a result, our findings capture overall travel time patterns rather than differences to MCH access between individual transport modes. Despite this, the study introduced a novel method to assess MCH accessibility in India, analyzing density, travel times, and APCs across 36 states/union territories and 735 districts. It compared rural and urban access to public and private MCHs, with validated geolocation strengthening the findings. Additionally, sensitivity analysis ensured the robustness of our findings.

### Future Directions

Future research should examine accessibility at finer geographic and demographic resolutions better to understand inequities in travel time and service reach. District- or state-specific analyses using higher-resolution road, traffic, and transport-mode data could help disaggregate motorized travel into public and private options and more accurately reflect real mobility patterns. Linking accessibility metrics to outcomes, such as maternal mortality, neonatal outcomes, emergency referrals, and surgical delays, would also strengthen validation efforts. Additionally, integrating facility readiness, service availability, and quality of care into accessibility models would provide a more holistic understanding of MCH access in India and guide more targeted policy interventions.

## 5. Conclusion

Median travel time to MCHs and Access Population Coverage (APC) are key tools for evaluating geographic accessibility and guiding policy. Our study revealed poor MCH access in Ladakh, the Andaman and Nicobar Islands, and the northeastern states of Arunachal, Nagaland, Mizoram, and Meghalaya. Rural areas showed significantly worse access than urban areas. Future research should examine accessibility for different demographics and consider the availability and quality of facilities in each MCH.

## Data Availability

All data produced in the present study are available upon reasonable request to the authors.
All data produced in the present work are contained in the manuscript.

## Declarations

### Ethics and Guidelines

**Consent to participate:** Not Applicable **Consent for publication:** Not Applicable **Ethical Approval:** Not Applicable

**Availability of data and materials:** The datasets used and/or analysed during the current study are available from the corresponding author on reasonable request.

**Competing interests:** Siddhesh Zadey represents the Association for Socially Applicable Research (ASAR) on the drafting committee of the Maharashtra State Mental Health Policy and the Global Alliance for Surgical, Obstetric, Trauma, and Anesthesia Care (G4 Alliance). He serves as the chair of the South Asia Working Group of the G4 Alliance. He is on the board of ASAR and the advisory board of Nivarana. He has previously received honoraria from Think Global Health and The Hindu. Other authors declare no competing interests.

**Funding:** None

## Authors’ contributions

***Study concept and design:*** Siddhesh Zadey

***Acquisition, analysis, or interpretation of data:*** Chaitanya Reddy, Harsh Thakkar, Aamir Miyajiwala, Sahil Kale, Ankit Raj, Siddhesh Zadey

***Drafting of the manuscript:*** Chaitanya Reddy, Varun Raj Passi

Critical revision of the manuscript for important intellectual content: ***All authors***

***Statistical analysis:*** Harsh Thakkar, Aamir Miyajiwala, Sahil Kale, Ankit Raj, Siddhesh Zadey

***Administrative, technical, or material support:*** Siddhesh Zadey

***Study supervision:*** Siddhesh Zadey

## Acknowledgments

Not Applicable

Clinical Trial Number: Not Applicable

## Notes

### Funding Statement

This study did not receive any funding.

### Summary of Updates

We added a district level correlation analysis to examine the relationship between geographic accessibility to MCHs and overall health system performance. Using the Universal Health Coverage (UHC) index as a reference indicator, we observed a consistent positive association with APC by motorized transport across all travel time thresholds. The association was strongest for access within 30 minutes, with a Spearman Rank correlation (R) of 0.308 and remained positive at 60 minutes (R = 0.275) 90 minutes (R = 0.218) and 120 minutes (R = 0.125) (eFigure 6D). Districts where a higher proportion of the population could reach MCHs by walking within shorter time limits tended to have better UHC index scores, with an R = 0.297 for 30 minutes and R = 0.316 for 60 minutes. The association was comparable to that observed for motorized travel and was strongest at shorter walking thresholds. This finding is important because walking access reflects local service availability and is less influenced by transport infrastructure. The consistency of results across both motorized and walking modes suggests that the accessibility metrics such as APC are robust and capture meaningful differences in health system access. These findings support the validity of APC as a meaningful measure of geographic accessibility that aligns with broader health system performance at the district level.

