## Supplementary figures and images for "Geospatial Modeling Study Assessing Population Level Accessibility to Medical College Hospitals in India"

### Supplementary Figures

#### Slide 1
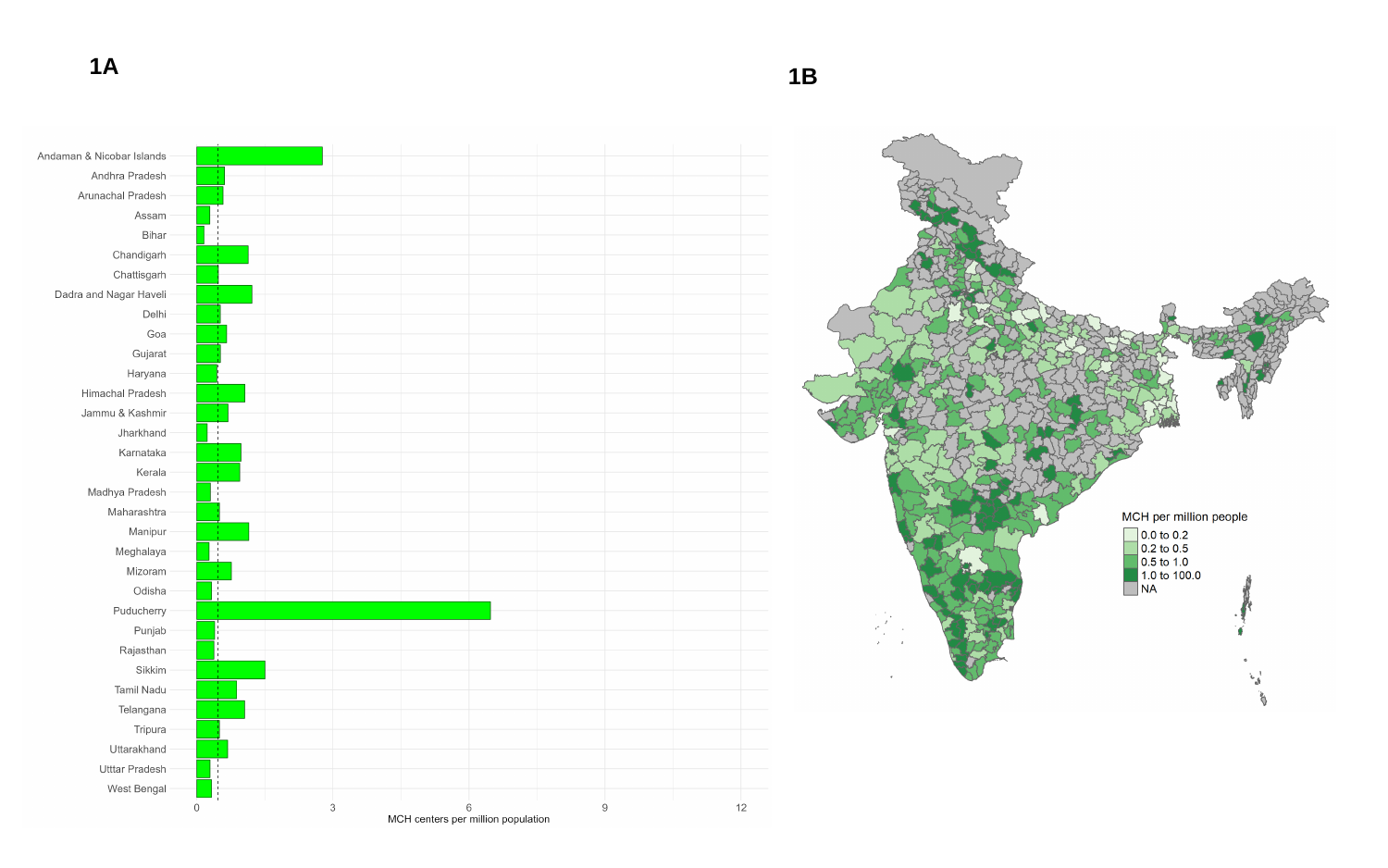

1A
1B

#### Slide 2
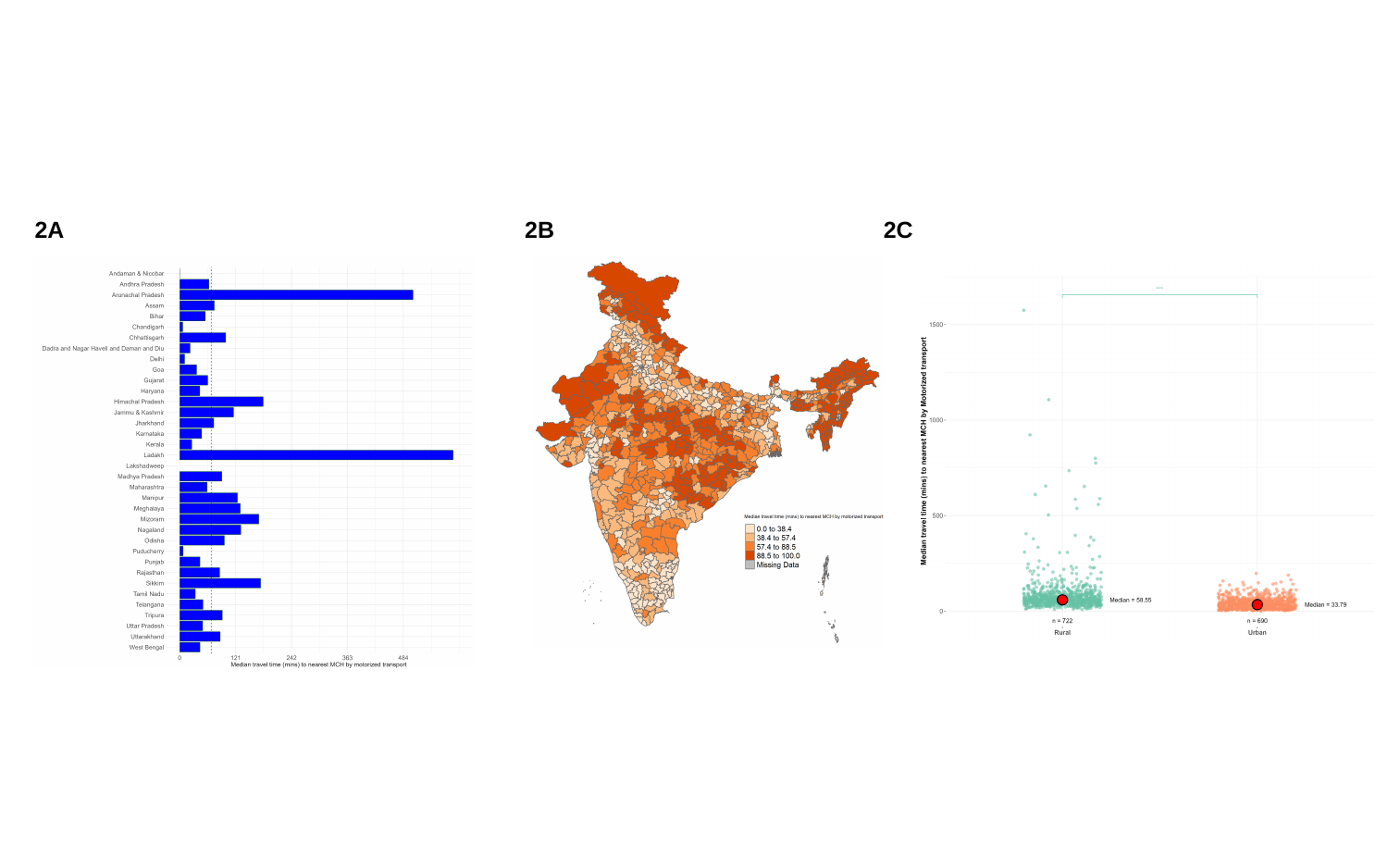

2A
2B
2C

#### Slide 3
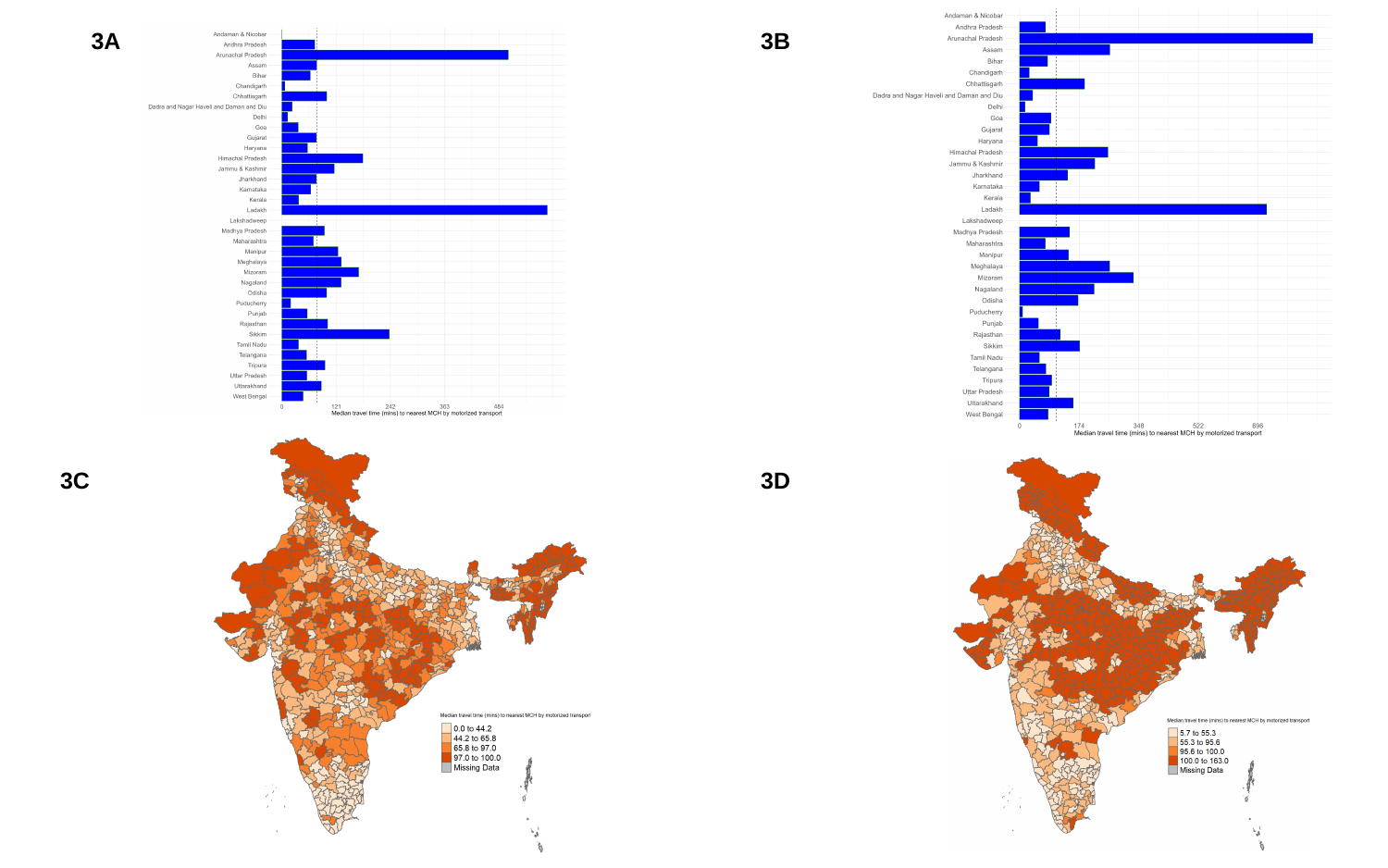

3A
3B
3C
3D

#### Slide 4
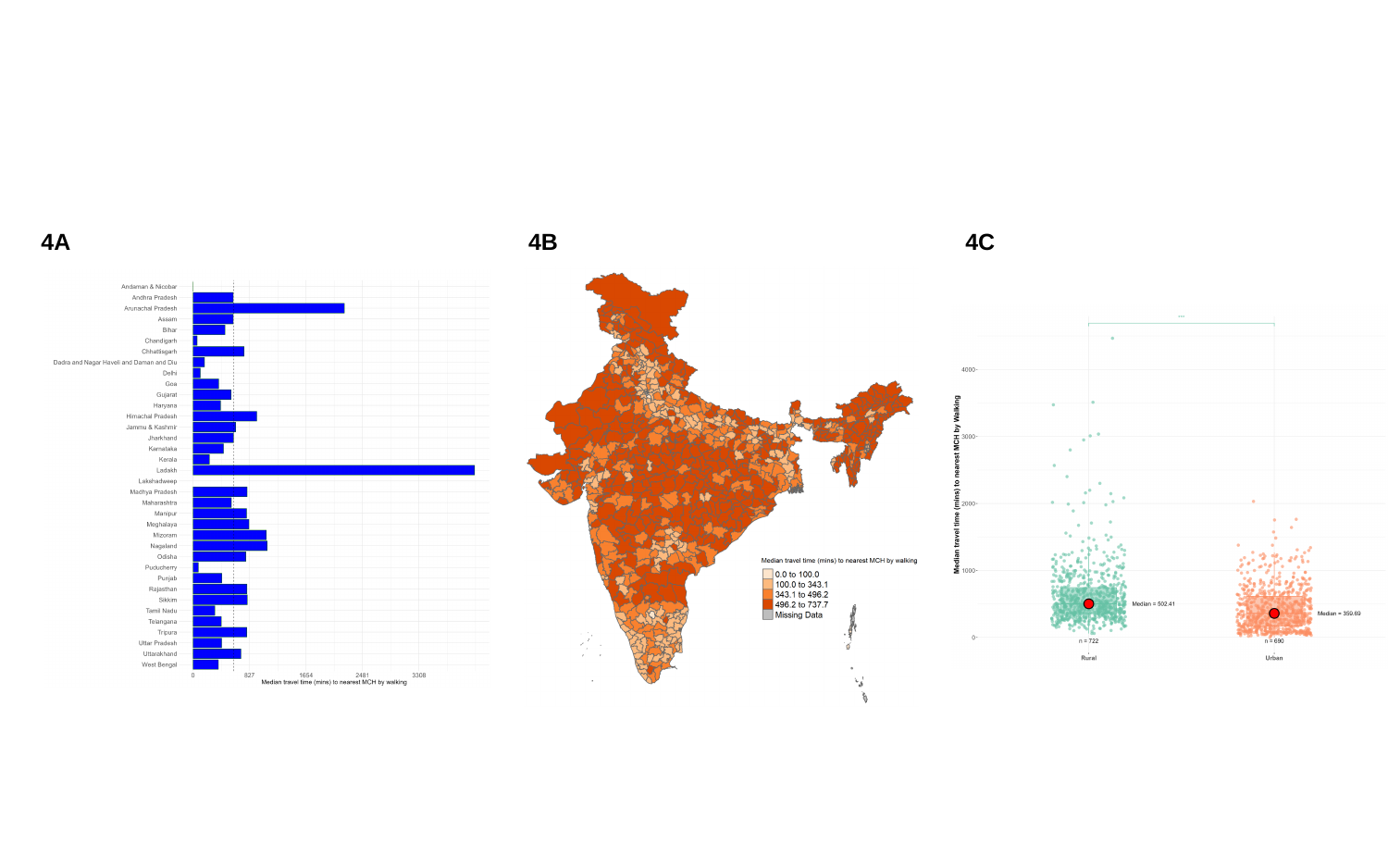

4A
4B
4C

#### Slide 5
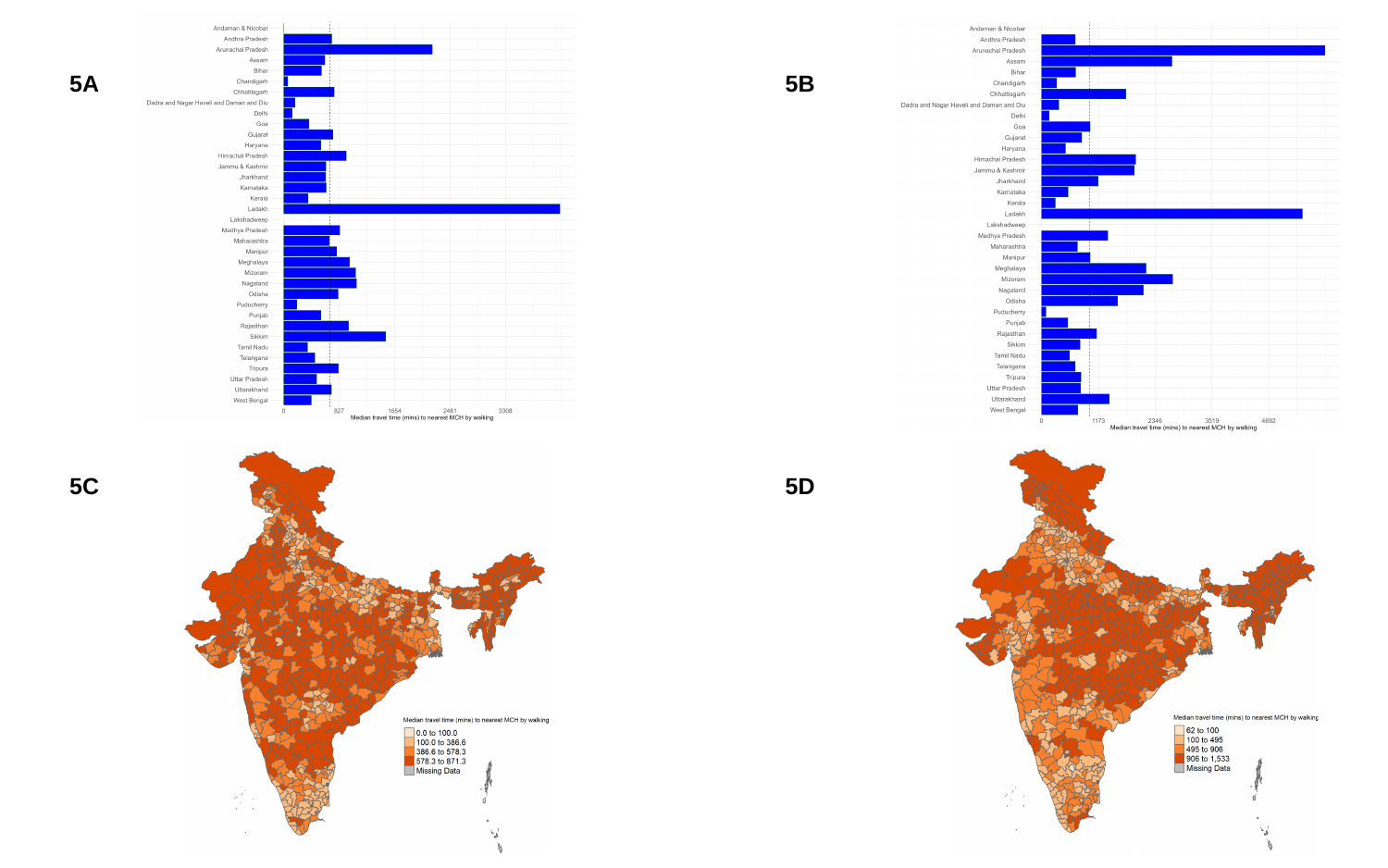

5A
5B
5C
5D

#### Slide 6
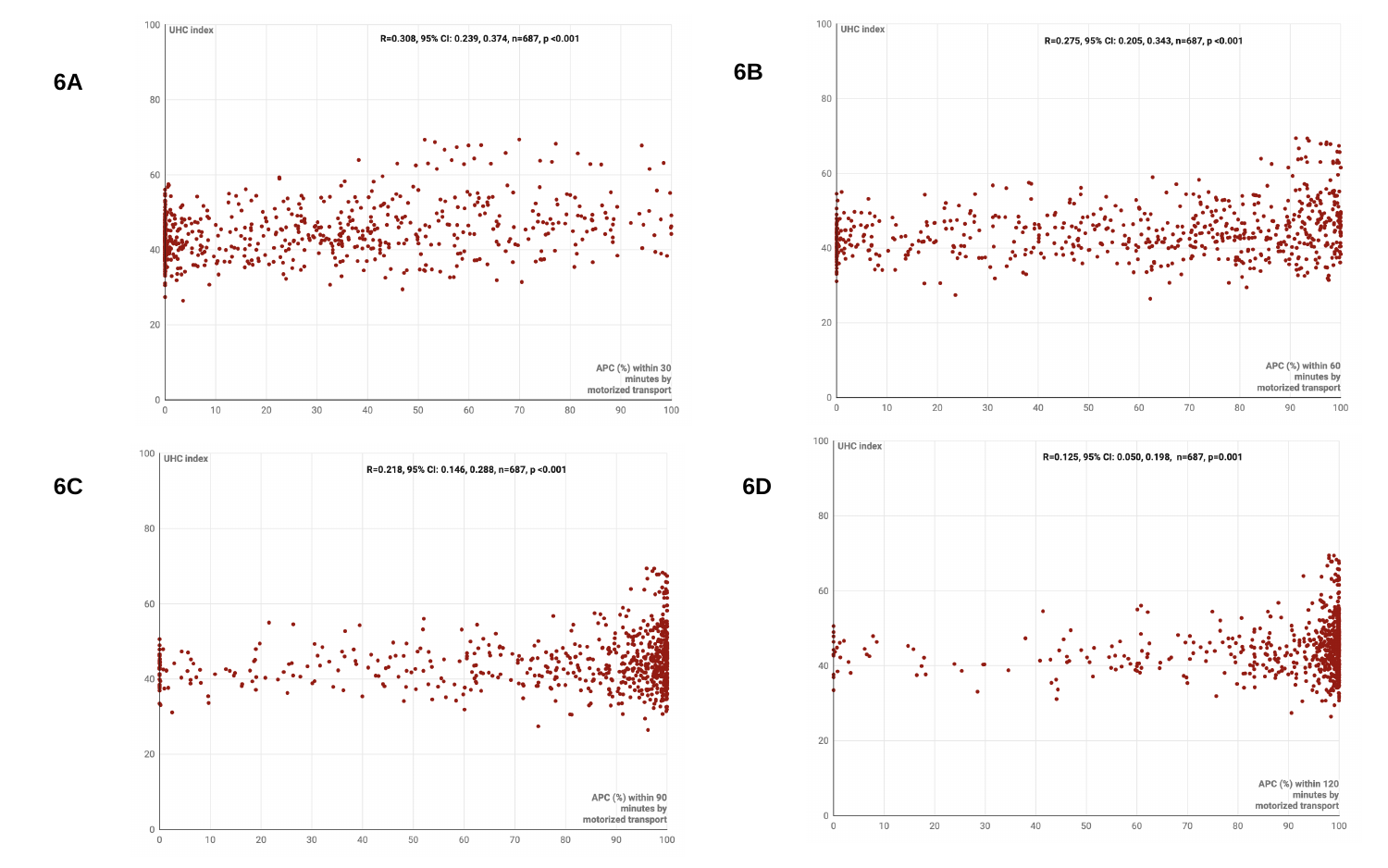

6B
6A
6C
6D

#### Slide 7
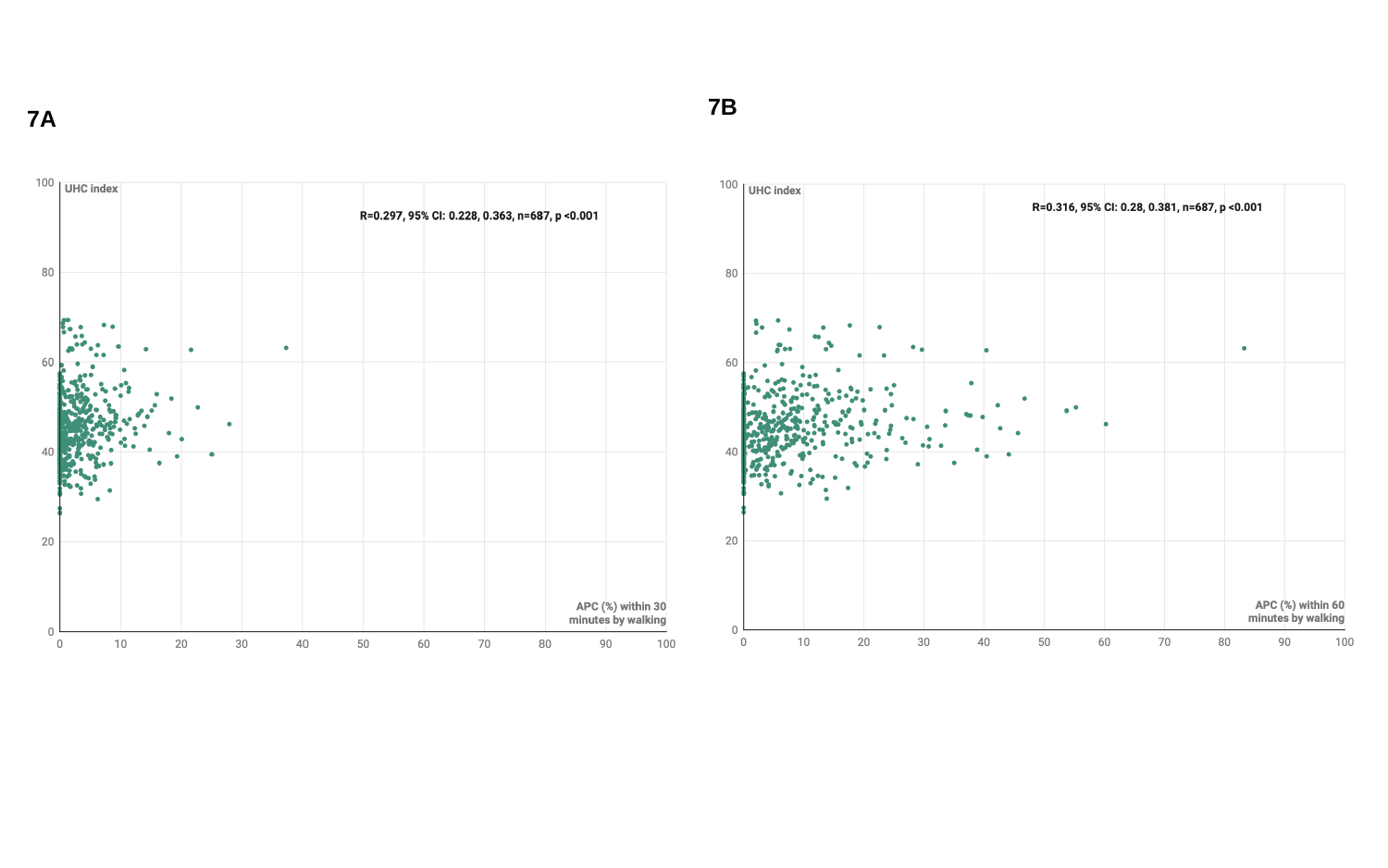

7B
7A
