## Supplementary Figure Captions for "Geospatial Modeling Study Assessing Population Level Accessibility to Medical College Hospitals in India"

**eFigure 1: MCH density per million for A. States and union territories and B. Districts. The states/UTs of Ladakh, Lakshadweep, and Nagaland are not mentioned as they have no MCHs. Districts having zero MCHs are marked as N/A.**

**eFigure 2: Median travel time to the nearest MCH by motorized transport for A. States and union territories. The black dotted line represents the national median travel time. B. Districts** **and C. Rural areas and urban areas.**

**eFigure 3: Median travel time to the nearest MCH by motorized transport for A. States and union territories with public MCHs. B. States and union territories with private MCHs**. **C. Districts with public MCHs and D. Districts with private MCHs.**

**eFigure 4: Median travel time to the nearest MCH by walking for A. States and union territories. The black dotted line represents the national median travel time. B. Districts** **and** **C. Rural areas and urban areas.**

**eFigure 5: Median travel time to the nearest MCH by walking for A. States and union territories with public MCHs. B. States and union territories with private MCHs**. **C. Districts with public MCHs and D. Districts with private MCHs.**

**eFigure 6: Scatter plots depicting bivariate distribution of UHC Index and APC values (%) for reaching nearest MCH by motorized transport at district levels within A. 30 minutes. B. 60 minutes. C. 90 minutes and D. 120 minutes. The R values represent the Spearman correlation coefficients. We used an alpha threshold of 0.05 for the associated p-values.**

**eFigure 7: Scatter plots depicting bivariate distribution of UHC Index and APC values (%) for reaching nearest MCH by walking at district levels within A. 30 minutes and B. 60 minutes. The R values represent the Spearman correlation coefficients. We used an alpha threshold of 0.05 for the associated p-values.**
